# Do nuclear magnetic resonance (NMR)-based metabolomics improve the prediction of pregnancy-related disorders?

**DOI:** 10.1101/2020.06.22.20134650

**Authors:** Nancy McBride, Sara L. White, Lucilla Poston, Diane Farrar, Jane West, Naveed Sattar, Scott M. Nelson, John Wright, Dan Mason, Matthew Suderman, Caroline Relton, Paul Yousefi, Deborah A Lawlor

**Affiliations:** MRC Integrative Epidemiology Unit at the University of Bristol, Bristol, UK; NIHR Bristol Biomedical Research Centre, University of Bristol, Bristol, UK; Population Health Sciences, University of Bristol, Bristol, UK; Department of Women and Children’s Health, Faculty of Life Sciences and Medicine, King’s College London, London, UK; Bradford Institute for Health Research, Bradford Teaching Hospitals NHS Foundation Trust, Bradford, UK; Cardiovascular and Medical Sciences, British Heart Foundation Glasgow, Cardiovascular Research Centre, University of Glasgow, Glasgow, UK; School of Medicine, University of Glasgow, Glasgow, UK

**Author notes:** Corresponding author: Nancy McBride,.

## Abstract

**Background:** Prediction of pregnancy-related disorders is mostly done based on established and easily measured risk factors. However, these measures are at best moderate at discriminating between high and low risk women. Recent advances in metabolomics may provide earlier and more accurate prediction of women at risk of pregnancy-related disorders.

**Methods and Findings:** We used data collected from women in the Born in Bradford (BiB; n=8,212) and UK Pregnancies Better Eating and Activity Trial (UPBEAT; n=859) studies to create and validate prediction models for pregnancy-related disorders. These were gestational diabetes mellitus (GDM), hypertensive disorders of pregnancy (HDP), small for gestational age (SGA), large for gestational age (LGA) and preterm birth (PTB). We used ten-fold cross-validation and penalised regression to create prediction models. We compared the predictive performance of 1) risk factors (maternal age, pregnancy smoking status, body mass index, ethnicity and parity) to 2) nuclear magnetic resonance-derived metabolites (N = 156 quantified metabolites, collected at 24-28 weeks gestation) and 3) risk factors and metabolites combined. The multi-ethnic BiB cohort was used for training and testing the models, with independent validation conducted in UPBEAT, a study of obese pregnant women of multiple ethnicities.

In BiB, discrimination for GDM, HDP, LGA and SGA was improved with the addition of metabolites to the risk factors only model. Risk factors area under the curve (AUC 95% confidence interval (CI)): GDM (0.69 (0.64, 0.73)), HDP (0.74 (0.70, 0.78)) and LGA (0.71 (0.66, 0.75)), and SGA (0.59 (0.56,0.63)). Combined AUC 95% (CI)): GDM (0.78 (0.74, 0.81)), HDP (0.76 (0.73, 0.79)) and LGA (0.75 (0.70, 0.79)), and SGA (0.66 (0.63,0.70)). For GDM, HDP, LGA, but not SGA, calibration was good for a combined risk factor and metabolite model. Prediction of PTB was poor for all models. Independent validation in UPBEAT at 24-28 weeks and 15-18 weeks gestation confirmed similar patterns of results, but AUC were attenuated. A key limitation was our inability to identify a large general pregnancy population for independent validation.

**Conclusions:** Our results suggest metabolomics combined with established risk factors improves prediction GDM, HDP and LGA, when compared to risk factors alone. They also highlight the difficulty of predicting PTB, with all models performing poorly.

**Author Summary:** *Background:*

- Current methods used to predict pregnancy-related disorders exhibit modest discrimination and calibration.
- Metabolomics may enable improved prediction of pregnancy-related disorders.

*Why Was This Study Done?:*

- We require tools to identify women with high-risk pregnancies earlier on, so that antenatal care can be more appropriately targeted at women who need it most and tailored to women’s needs and to facilitate early intervention.
- It has been suggested that metabolomic markers might improve prediction of future pregnancy-related disorders. Previous studies tend to be small and rarely undertake external validation.

*What Did the Researchers Do and Find?:*

- Using BiB (8,212 pregnant women of multiple ethnicities), we created prediction models, using established risk factors and 156 NMR-derived metabolites, for five pregnancy-related disorders. These were gestational diabetes mellitus (GDM), hypertensive disorders of pregnancy (HDP), small for gestational age (SGA), large for gestational age (LGA) and preterm birth (PTB). We sought external validation in UPBEAT (859 obese pregnant women).
- We compared the predictive discrimination (area under the curve - AUC) and calibration (calibration slopes) of the models. The prediction models we compared were 1) established risk factors (pregnancy smoking, maternal age, body mass index (BMI), maternal ethnicity and parity) 2) NMR-derived metabolites measured in the second trimester and 3) a combined model of risk factors and metabolites.
- Inclusion of metabolites with risk factors improved prediction of GDM, HDP, LGA and SGA in BiB. Prediction of PTB was poor with all models. Result patterns were similar in validation using UPBEAT, particularly for GDM and HDP, but AUC were attenuated.

*What Do These Findings Mean?:*

- These findings indicate that combining current risk factor and metabolomic data could improve the prediction of GDM, HDP, LGA and SGA. These findings need to be validated in larger, general populations of pregnant women.

## Introduction

Around 40% of all pregnancies are complicated by one or more of gestational diabetes (GDM), hypertensive disorders of pregnancy (HDP), small or large for gestational age (SGA, LGA) and preterm birth (PTB). These pregnancy-related disorders have adverse short- and long-term consequences for mother and child ^1-7^. Established risk factors for pregnancy-related disorders include pregnancy smoking, maternal age, body mass index (BMI), maternal ethnicity and parity ^6, 8-12^. However, a large proportion of disorders occur in women without any known risk factors. Current identification of women who are ‘high-risk’ uses clinical screening of these risk factors, sometimes in combination with early pregnancy measures of glucose for GDM ^13^, blood pressure for PE ^6^, ultrasound for SGA and LGA ^14^ and cervical length measurement/fetal fibronectin for (PTB) ^15^. However, whilst glucose measures in early pregnancy can identify women with undiagnosed existing diabetes, neither it, nor established risk factors in early pregnancy, predict GDM risk accurately ^16^. Ultrasound has poor consistency, is prone to human error and often fails to identify SGA or LGA babies until very late in pregnancy ^17^. Cervical length and fetal fibronectin have improved the prediction of PTB, but are invasive and only predict ‘imminent’ preterm birth in women where this is suspected ^15^.

These pregnancy-related disorders often co-occur, with women with GDM more likely to have pregnancies complicated by hypertension or pre-eclampsia (PE), and their offspring being born LGA ^2^. Similarly, women with HDP are more likely to have their offspring born SGA or preterm ^5^. However, most research focuses on single outcomes. This multimorbidity should be addressed to see if a common prediction tool, or a tool with an overlap of variables can be developed for predicting global risk of several pregnancy-related disorders. It may also enable identification of women likely to have a healthy pregnancy ^18 19, 20^

Metabolites might improve prediction of pregnancy-related disorders. Metabolite levels are known to change markedly during pregnancy ^21, 22^, associate with cardio-metabolic outcomes (known correlates of pregnancy-related disorders) ^18^, and with pregnancy-related disorders in some studies ^23^. Most studies exploring the value of metabolomics in predicting pregnancy-related disorders have focused on GDM, PE or SGA. The most notable omics predictor that has been identified to date is soluble fms-like tyrosine kinase 1 (sFlt-1) and placental growth factor (P1GF) ratio for predicting PE. sFlt-1:P1GF is an accurate predictor of PE in both low and high-risk pregnant women ^24^. With respect to metabolite prediction, two studies reported excellent predictive discrimination for SGA (AUC > 0.90) - one study which developed a metabolomic model of five metabolites ^25^ and another of 19 metabolites ^26^. However, these were based on small samples of 83 and 8 women, respectively. Similarly, a study reported that a panel of four mass-spectrometry derived metabolites could predict spontaneous PTB with a partial AUC (i.e. an alternative to AUC, whereby only the regions of ROC space where data are observed are included) of 12.6 in 105 women ^27^. These studies did not compare their models to existing risk factors or undertake external validation. A systematic review of metabolomic prediction of SGA identified 15 studies ^28^. Of these, only three were designed for prediction purposes and provided any metric of prediction. Two of these three had sample sizes of 80 and 83 women. None of them sought external validation. For GDM, nuclear magnetic resonance (NMR)-derived metabolites have been found to distinguish between women who did and did not go on to develop GDM, when looked at in early pregnancy. However, discrimination did not improve when added to a risk prediction model of candidate biomarkers^29^.

A recent collaboration between the Pregnancy Outcomes Prediction study (POPs) and the Born in Bradford (BiB) cohort (the latter used as external validation) using mass-spectrometry metabolomics (>1100 semi-quantified untargeted metabolites) has shown that 4-hydroxyglutamate improves prediction of PE over risk factors alone ^30^. The same collaboration found that sFlt-1:P1GF and a ratio of combining four metabolites (1-(1-enyl-stearoyl)-2-oleoyl-GPC, 1,5-anhydroglucitol,5α-androstan-3α,17α-diol disulfate and N1,N12-diacetylspermine) is a better predictor of fetal growth restriction than sFlt-1:P1GF combined with risk factors ^31^.

In this study, we aim to see whether NMR-derived metabolites can improve the prediction of pregnancy-related disorders, over and above established risk factors (pregnancy smoking, maternal age, BMI, maternal ethnicity, and parity). We focus on the prediction of five common pregnancy-related disorders: GDM, HDP, SGA, LGA and PTB. We used two samples, 1) women in the BiB cohort, used for training and testing the prediction models and 2) obese pregnant women (BMI ≥ 30kg/m^2^) in the UPBEAT study, used for external validation of the prediction models.

## Methods

### Participants

We used data from the BiB study, a population-based prospective birth cohort that recruited 12,453 women who had 13,776 pregnancies. Full details of the study methodology were reported previously ^32^. In brief, most women were recruited at their oral glucose tolerance test (OGTT) at approximately 26–28 weeks gestation, which was offered to all women booked for delivery at Bradford Royal Infirmary at the time of recruitment. Eligible women had an expected delivery between March 2007 and December 2010. Ethical approval for the study was granted by the Bradford National Health Service Research Ethics Committee (ref 06/Q1202/48). The UPBEAT study was a multicentre randomised control trial (RCT) which recruited 1,555 obese pregnant women (BMI ≥ 30kg/m^2^) between 15-18+6 weeks gestation, at eight centres across the UK ^33^. UPBEAT is registered with Current Controlled Trials (ISRCTN89971375) and approvals were obtained from the UK research ethics committee (ref 09/H0802/5). Local Research and Development departments in participating centres approved participation of their respective centres. All women in both studies provided written informed consent. Figure 1 illustrates the flow of participants. To be eligible for inclusion in the analysis, all women had to have a fasting pregnancy serum sample (used for NMR metabolome profiling), information on all established risk factor predictors and all pregnancy-related disorders. This resulted in 8,212 BiB women and 859 UPBEAT women being included. We use UPBEAT here as a cohort study, including both arms of the trial combined and adjusting for which arm they were allocated to. UPBEAT is a RCT looking at the effect of a tailored lifestyle intervention aimed at improving diet and physical activity ^33^. The UPBEAT intervention did not influence the primary outcome of GDM, or any of the pregnancy-related disorders explored here ^34^. It did influence change in several lipids, fatty acids and some amino acids from the NMR platform used here ^34^.

**Figure 1:**
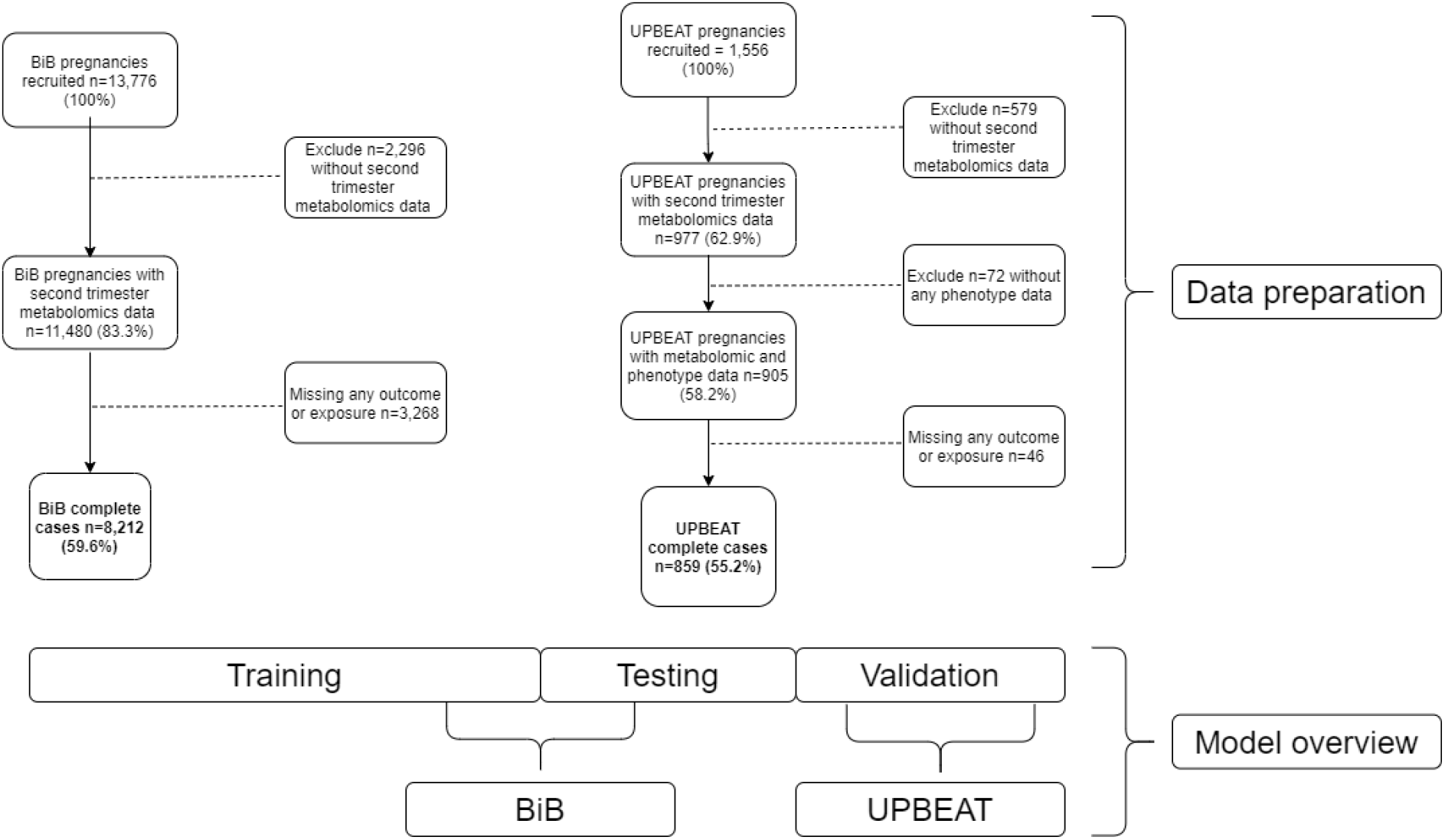
Data overview: flow of participants (above) in Born in Bradford (BiB) cohort (top left) and UK Pregnancies Better Eating Activity Trial (UPBEAT) randomised control trial (RCT) (top right) to generate the final sample for analysis. Model overview: sample split for model selection (below middle) Abbreviations: BMI, body mass index; GDM, gestational diabetes; HDP, hypertensive disorder of pregnancy, SGA, small for gestational age; LGA, large for gestational age; PTB, preterm birth.

### Metabolomic profiling

In both studies comprehensive metabolomic profiling was performed using high throughput targeted NMR platform (Nightingale Health (http://www.computationalmedicine.fi/), (Helsinki, Finland) run either at the University of Bristol (BiB) or Nightingale Health (under its previous name of Brainshake) (UPBEAT). Of the 13,776 pregnancies in the BiB cohort, 11,476 pregnancies had a fasting serum sample taken at a single timepoint, between 24-28 weeks gestation. In UPBEAT, NMR profiling was conducted at three time points during pregnancy 15-18+6 weeks, 27-28+6 weeks, 34-36 weeks gestation ^33^. We used the 27-28+6-week timepoint for our main analyses because it matched the gestational age at which BiB samples were taken for metabolomic profiling and, like BiB, were fasting samples. The NMR platform quantified 156 metabolic traits. The targeted metabolic traits measured by the platform represent a broad molecular signature of systemic metabolism including routine lipids, lipoprotein subclass profiling, fatty acid composition, and several low-molecular metabolites, including amino acids, ketone bodies and gluconeogenesis-related metabolites, mostly in molar concentration units. A full list of all the traits is provided in Table S1. The NMR platform has been applied in various large-scale epidemiological studies, with detailed protocol and quality control information being previously published ^35, 36^.

### Maternal pregnancy measurements

For all outcomes we compared the predictive ability of the metabolomic measures in relation to a set of common predictors that are routinely used in antenatal care to risk stratify women: maternal age, early-pregnancy/recruitment BMI, parity, ethnicity and smoking during pregnancy. This information was collected during recruitment or extracted from clinical records in both studies. All pregnancies included in this study were singleton pregnancies. In both studies data on parity were extracted from the first antenatal clinic records (around 12-weeks of gestation) and categorized as having experienced one or more previous pregnancy ≥ 24 weeks gestation, or no previous pregnancy. Ethnicity was self-reported or obtained from primary care medical records. It was categorised using UK Office of National Statistics criteria: 1) White European (‘White British’ or ‘White European’); 2) South Asian (‘Pakistani’, ‘Indian’ or ‘Bangladeshi’); 3) Caribbean or African (‘Afro-Caribbean’ or ‘African’) or 4) Other. Information on maternal age and smoking were obtained at recruitment (24-28 weeks gestation in BiB and 15-18+6 weeks in UPBEAT) via researcher interview. Smoking was dichotomised as any smoking during pregnancy. In BiB, weight was extracted from the first antenatal clinic (∼12-weeks) and height measured at recruitment. In UPBEAT, weight and height were measured at recruitment (15-18+6 weeks).

We examined predictive discrimination for five pregnancy-related disorders: GDM, HDP, SGA, LGA and PTB. In BiB all blood pressure measures and proteinuria measurements taken at any time during pregnancy were extracted from medical records ^1^. In UPBEAT these measures were taken at the participating centres. In both studies, gestational hypertension was defined as new onset of elevated blood pressure (systolic blood pressure ≥140 mmHg or greater, and/or diastolic blood pressure ≥90 mmHg or greater) after 20 weeks’ gestation on two or more occasions. PE was defined as gestational hypertension plus clinically significant proteinuria, defined as 1 or greater ‘+’ on the reagent strip reading (equivalent to 30mg/mmol) or greater on spot urine protein/creatinine ratio). We *a priori* decided that there were too few cases in BiB to examine prediction of PE separately from gestational hypertension so combined these to generate the ‘hypertensive disorder of pregnancy’ variable used in this study. All women in BiB and UPBEAT were offered a 75-g OGTT at 27-28 weeks of gestation. In BiB, fasting and 2hr post-load samples were collected and analysed; in UPBEAT, fasting, 1hr and 2hr glucose were collected and analysed. In BiB, GDM was defined according to modified World Health Organization (WHO) criteria operating at the time of the study; fasting glucose ≥ 6.1 mmol/L or 2hr post-load glucose ≥ 7.8 mmol/l ^3^. In UPBEAT, GDM was defined according to the guidelines recommended by the International Association of Diabetes and Pregnancy Study Groups (IADPSG); fasting glucose ≥ 5.1 mmol/L, 1-hour glucose ≥ 10.0 mmol/L or higher, 2hr venous glucose of ≥ 8.5 mmol/L) ^37^. In both studies, UK WHO fetal growth charts were used as the external standard for generating gestational age and sex standardised birthweight percentiles. SGA was defined as <10^th^ percentile and LGA as >90^th^ percentile. In both studies PTB was defined as delivery before 37 completed weeks.

### Statistical analysis

#### General approach

We developed three prediction models for each pregnancy-related disorder: (i) established risk factors (maternal age, early-pregnancy/recruitment BMI, parity, ethnicity and smoking during pregnancy); (ii) NMR metabolites (156 metabolite traits) and (iii) combined risk factor and metabolomics predictors. Glucose was excluded from the metabolite prediction models for GDM because the samples had been taken at the OGTT and used to diagnose GDM. All three models were developed in a random subset of 75% of BiB (training set), and discrimination and calibration assessed in the remaining 25% of BiB (testing set). External validation in UPBEAT was undertaken by assessing the performance of the models developed in the BiB training subset.

Having developed models for each outcome separately, we explored the extent to which these were consistent across outcomes based on the variables included in BiB. We also explored discrimination of models developed for one outcome with other outcomes (details in ‘Sensitivity analyses’, below). This was done to assess the potential of having just one or a small number of models to predict all (or several) outcomes.

#### Model selection

We performed ten-fold cross-validation and penalised regression using the *caret* package in R version 3.5.1 ^38^. To construct a model in the training subset using elastic net, an optimal lambda parameter must first be selected. This is done by applying ten-fold cross validation to the training subset with a variety of lambda values. The lambda with the best cross-validated performance is then used to apply elastic net to the training set to obtain a final predictive model. The performance of this model is internally validated by applying it to the testing subset. This process is more robust than doing just one (training and testing) analysis ^39^. Penalised regression is a method for selecting which variables remain in the prediction model, variables whose coefficients are closer to the null are penalised (shrunk to zero) ^40-42^. We used optimal values of alpha and lambda (weights used in penalising) that minimize residual variance and hence maximise prediction. These cross-validation analyses were undertaken in a randomly selected 75% subset of the BiB cohort and then internal validation performed on the remaining 25%.

#### External validation

We were unable to identify an independent study with relevant metabolomic data in a general population of pregnant women for external validation. We therefore undertook external validation in a population of obese pregnant women (UPBEAT).

#### Assessing model discrimination and calibration for prediction of pregnancy outcomes

We assessed model discrimination using AUC, ranging from no discriminative ability (0.5) to perfect discriminative ability (1). We assessed calibration (the extent to which our model predicted probability of outcomes matched observed risk) using calibration slopes.

#### Sensitivity analysis

To explore whether the different definition of GDM in BiB and UPBEAT influenced results we estimated the AUC for our GDM model using the same OGTT criteria as those applied to BiB. We used women in UPBEAT’s individual glucose measurements to define as GDM using the criteria; fasting glucose ≥ 6.1 mmol/L or 2hr post-load glucose ≥ 7.8 mmol/l.

As our external validation sample was only in obese pregnant women, we were concerned that any failure to validate might be due to differences in BMI distribution between BiB and UPBEAT. To explore this, we compared the association of BMI with the five pregnancy-related disorders in 1) BiB, 2) in women in BiB with a BMI ≥ 30kg/m^2^ and 3) UPBEAT (where all women had BMI ≥ 30kg/m^2^). This would enable us to see if there was any evidence that BMI relates differently to the outcome when only obese women were included.

To evaluate whether we could use one model to predict more than one pregnancy-related disorder, we estimated the AUC for other outcomes using the models trained and tested in BiB that had an AUC >=0.6 for their specific outcome (e.g. we estimated the AUC for predicting HDP, SGA, LGA using the GDM models).

In the main analyses, we used the 27-28+6-week UPBEAT timepoint for the validation to match our discovery sample. We repeated analysis in UPBEAT using the earliest timepoint (15-18+6 weeks’ gestation) of metabolite measurements and explored the correlation between the 15-18+6 - and 27-28+6-week measures.

We examined prediction of spontaneous PTB, defined as those who had given birth before 37 weeks, with natural onset of labour (no medical or surgical induction). As there were only 15 spontaneous PTB in UPBEAT we did not seek to replicate the model that was trained and tested in BiB (n=260 spontaneous preterm).

## Results

Distributions of age, smoking, parity, HDP and PTB were broadly similar between the two cohorts. Differences in ethnicity reflected the sampling frame for each study. Other notable differences reflected the selection of only obese women in UPBEAT. They had higher mean BMI, and higher prevalence of GDM and LGA, but lower prevalence of SGA. The higher prevalence of GDM also reflects the different diagnostic criteria used in the two studies. Proportions remained higher in UPBEAT when the same criteria used in BiB were applied, but with a smaller difference between the two studies.

Table 1 shows the characteristics of the women in BiB and UPBEAT.

**Table 1.**
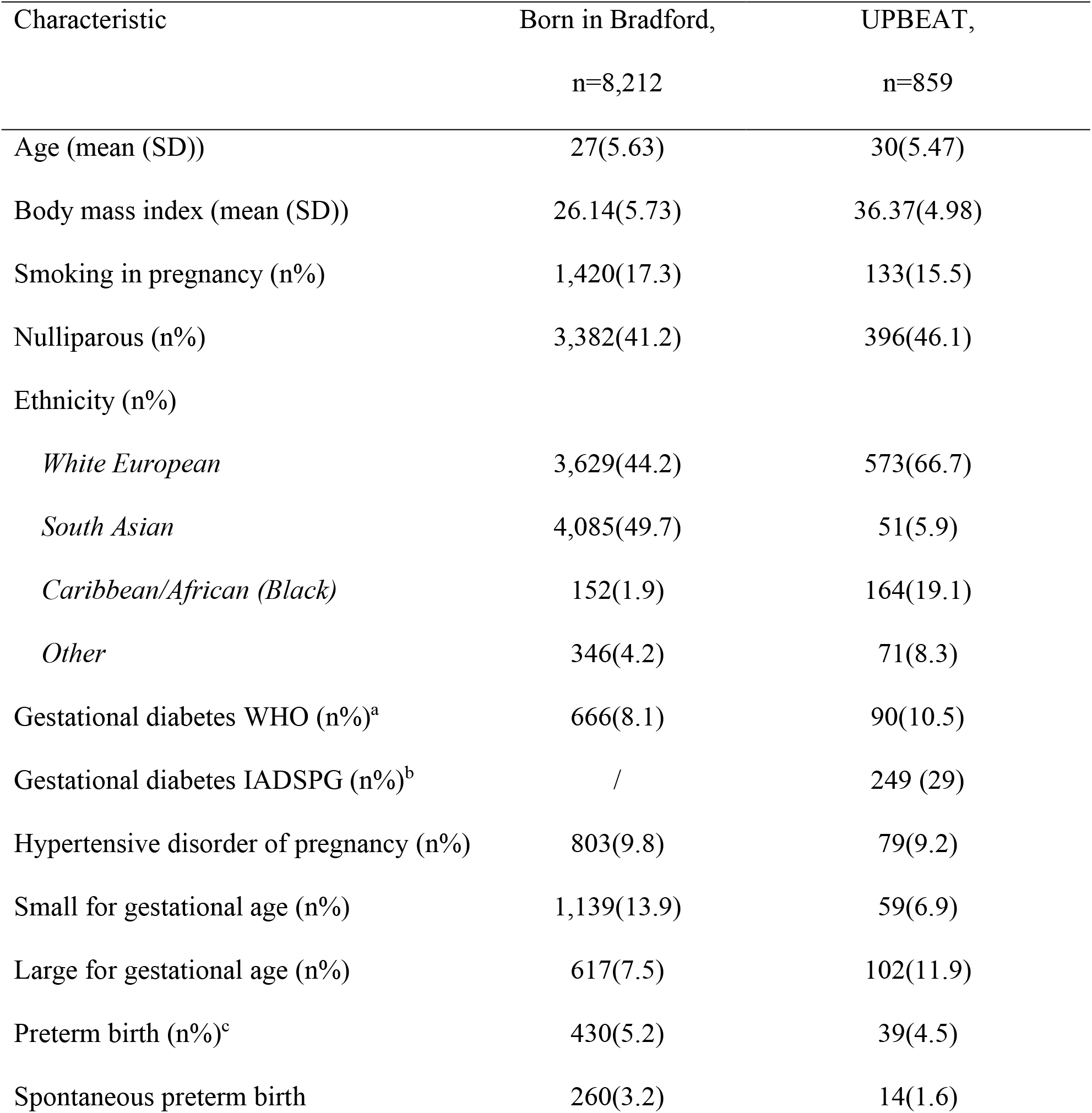
Data are expressed as mean (SD) or n (%) as appropriate. Data were 100% complete. Maternal age and weight/height (used to calculate body mass index (BMI)) were measured at recruitment. Smoking was defined as any smoking during pregnancy. Parity defined as this pregnancy being their first child (nulliparous) or having pr eviously giving birth (multiparous). Ethnicity was based on self-report. ^a^ Gestational diabetes was diagnosed in B orn in Bradford according to modified World Health Organization (WHO) criteria operating at the time of the st udy. ^b^ In UPBEAT, gestational diabetes was defined according to the guidelines recommended by the Internatio nal Association of Diabetes and Pregnancy Study Groups (IADSPG). We conducted a sensitivity analysis using t he WHO criteria in UPBEAT to check differences were not due to different GDM criteria. ^c^ Preterm birth includ es spontaneous and iatrogenic preterm birth (birth <37 weeks gestation).

| Characteristic | Born in Bradford,<br>n=8,212 | UPBEAT,<br>n=859 |
| --- | --- | --- |
| Age (mean (SD)) | 27(5.63) | 30(5.47) |
| Body mass index (mean (SD)) | 26.14(5.73) | 36.37(4.98) |
| Smoking in pregnancy (n%) | 1,420(17.3) | 133(15.5) |
| Nulliparous (n%) | 3,382(41.2) | 396(46.1) |
| Ethnicity (n%) |  |  |
| *White European* | 3,629(44.2) | 573(66.7) |
| *South Asian* | 4,085(49.7) | 51(5.9) |
| *Caribbean/African (Black)* | 152(1.9) | 164(19.1) |
| *Other* | 346(4.2) | 71(8.3) |
| Gestational diabetes WHO (n%)<sup>a</sup> | 666(8.1) | 90(10.5) |
| Gestational diabetes IADSPG (n%)<sup>b</sup> | / | 249 (29) |
| Hypertensive disorder of pregnancy (n%) | 803(9.8) | 79(9.2) |
| Small for gestational age (n%) | 1,139(13.9) | 59(6.9) |
| Large for gestational age (n%) | 617(7.5) | 102(11.9) |
| Preterm birth (n%)<sup>c</sup> | 430(5.2) | 39(4.5) |
| Spontaneous preterm birth | 260(3.2) | 14(1.6) |

### Variables included in the final models for each outcome and overlap between these

Table 2 shows the number of predictors retained in each model during model training in BiB. A full list of the predictors retained in any of the prediction models can be found in Tables S2-S4.

**Table 2.** Number of predictors retained in each model developed and tested in BiB from total possible (n(%)). Percentages are rounded to the nearest whole number.

| Outcome | Model (retained predictors/total number of variables possible [%]) |
| --- | --- |
| Gestational diabetes | Risk factor (5/5 [100%]) |
|  | Metabolite (140/156 [90%]) |
|  | Combined (152/161 [94%]) |
| Hypertensive disorder of pregnancy | Risk factor (4/5 [80%]) |
|  | Metabolite (50/156 [32%]) |
|  | Combined (38/161 [24%]) |
| Small for gestational age | Risk factor (4/5 [80%]) |
|  | Metabolite (86/156 [55%]) |
|  | Combined (101/161 [63%]) |
| Large for gestational age | Risk factor (4/5 [80%]) |
|  | Metabolite (65/156 [42%]) |
|  | Combined (56/161 [35%]) |
| Preterm birth | Risk factor (4/5 [80%]) |
|  | Metabolite (19/156 [12%]) |
|  | Combined (18/161 [11%]) |

Of the total 161 variables included in the combined model, most (94%) were retained in the GD model and least (11%) in the PTB model. At least 4, of the 5, established risk factors were retained in the combined models for all outcomes. The predominant metabolite classes retained in GDM, SGA and LGA outcomes were triglycerides, monounsaturated fatty acids, and apolipoproteins.

Only ten predictors were common across all models (Table S5). These were BMI, parity, smoking, ethnicity, creatinine, phenylalanine, isoleucine, glycine, valine, and glycerol.

### Model discrimination and calibration

Figure 2 shows the AUC for all three models with all outcomes in BiB (triangles) and UPBEAT (circles).

**Figure 2.**
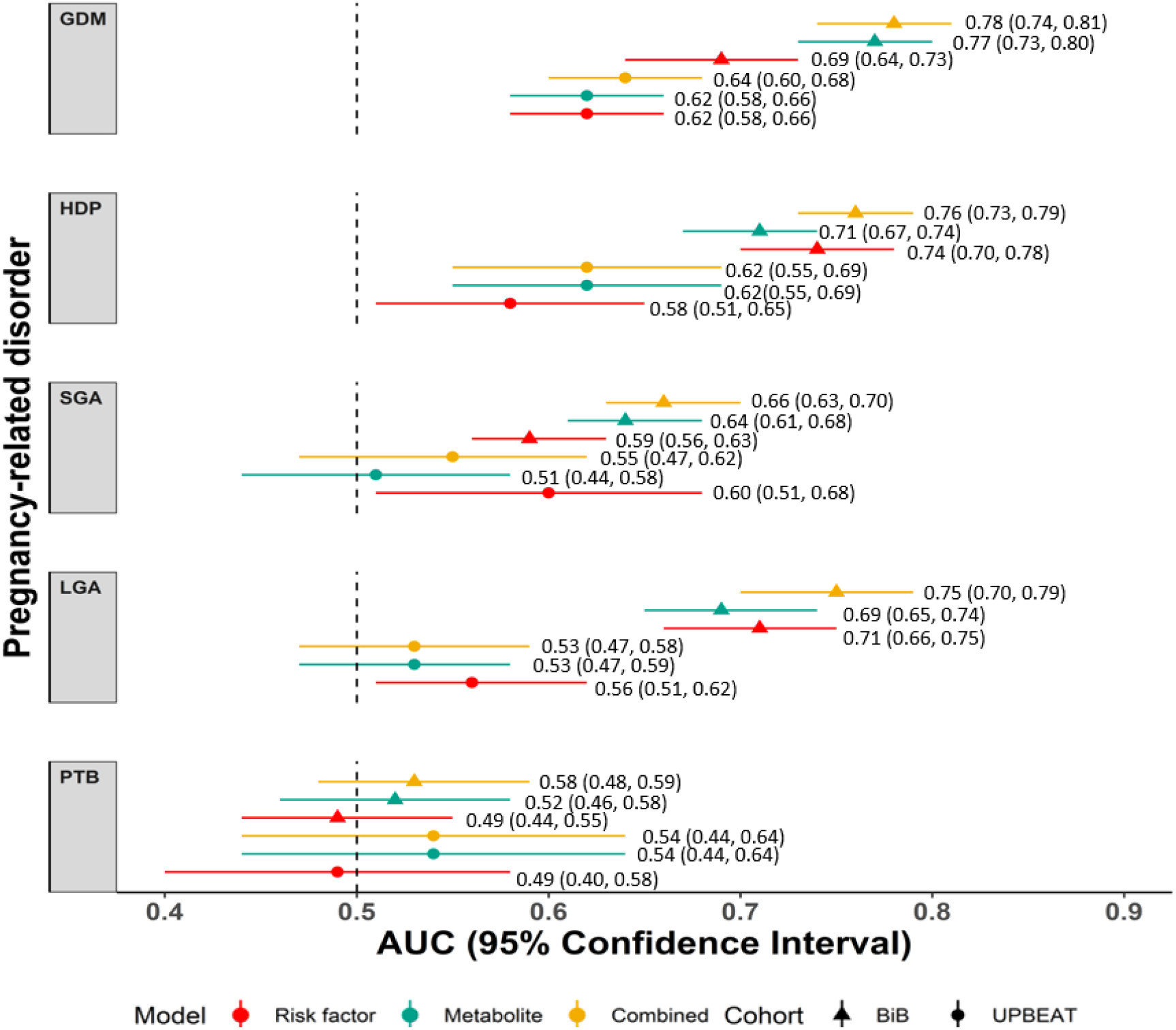
Predictive discrimination of models for each outcome. AUC and 95% confidence intervals are shown for established risk factor prediction models (red), metabolite models (green) and combined risk factor and metabolite models (yellow) in Born in Bradford (BiB) (triangles) and the UK Pregnancy Better Eating Activity Trial (UPBEAT) (circles). Abbreviations: GD, gestational diabetes; HDP, hypertensive disorders of pregnancy; SGA, small for gestational age; LGA, large for gestational age; PTB, preterm birth (iatrogenic or spontaneous) (Table S6).

In BiB, discrimination for GD, HDP and LGA was good (**Figure 2**, range of AUC for all models across these three outcomes 0.69 to 0.78) for all models and improved with the addition of metabolites to the risk factors only model, particularly for GDM (difference in AUC (95%CI): 0.09 (0.08, 0.10), 0.02 (0.03, 0.01) and 0.04 (0.04, 0.03)), respectively for GD, HDP and LGA). Modest discrimination for the SGA risk factors only model (AUC (95%CI) 0.59 (0.56-0.63)) improved when metabolites were added (AUC (95% CI) 0.66 (0.63,0.70)). For PTB discrimination was poor in all models (AUC∼0.5).

We evaluated calibration of the models which had performed well: GDM, HDP and LGA in BiB (Figures 3-5). As the intercepts on the slopes show, calibration is good for GD and LGA, but with some overestimation of GD and underestimation of LGA compared with the observed incidence. The combined model for HDP had the best calibration.

**Figure 3.**
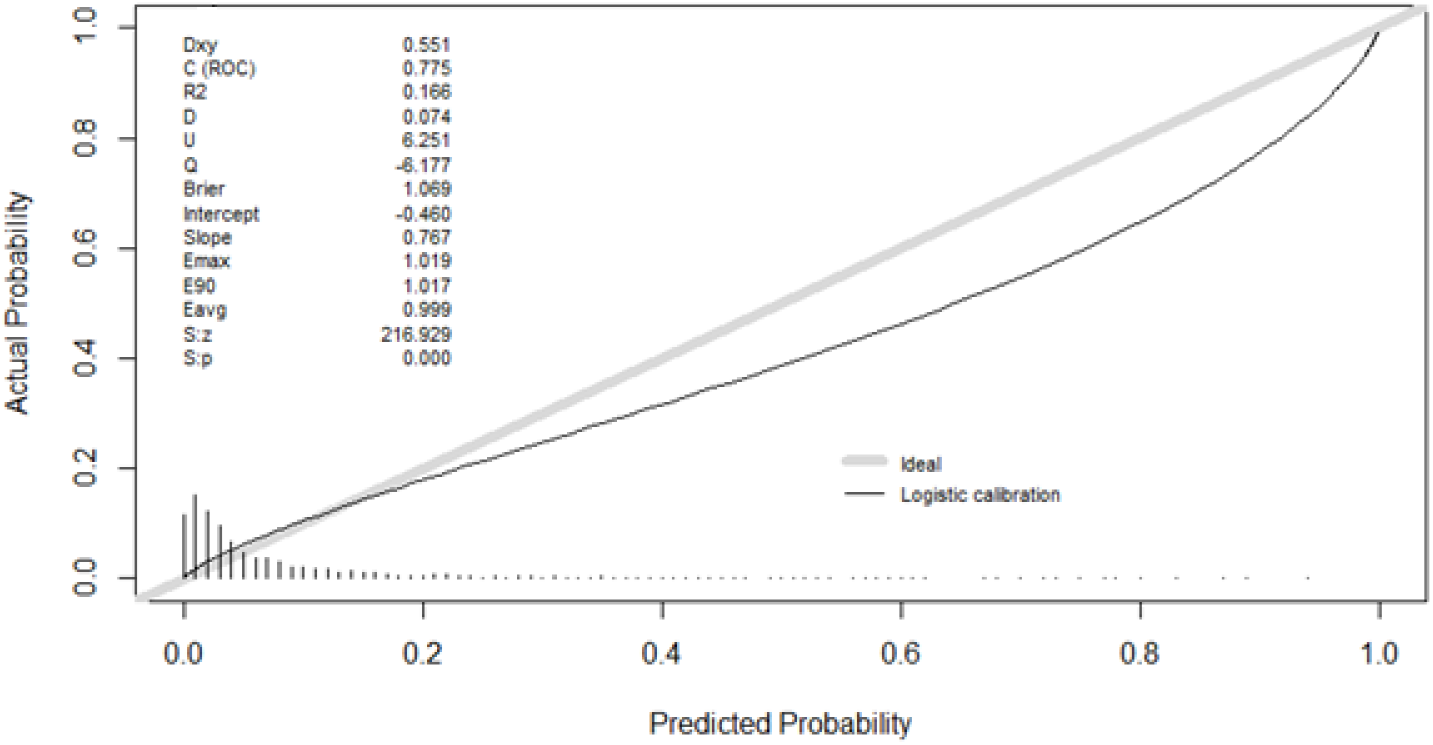
Calibration of combined model tested in BiB.

Figure 3 shows the calibration slope for the combined model for GD.

Figure 4 shows the calibration slope for the combined model for HDP.

**Figure 4.**
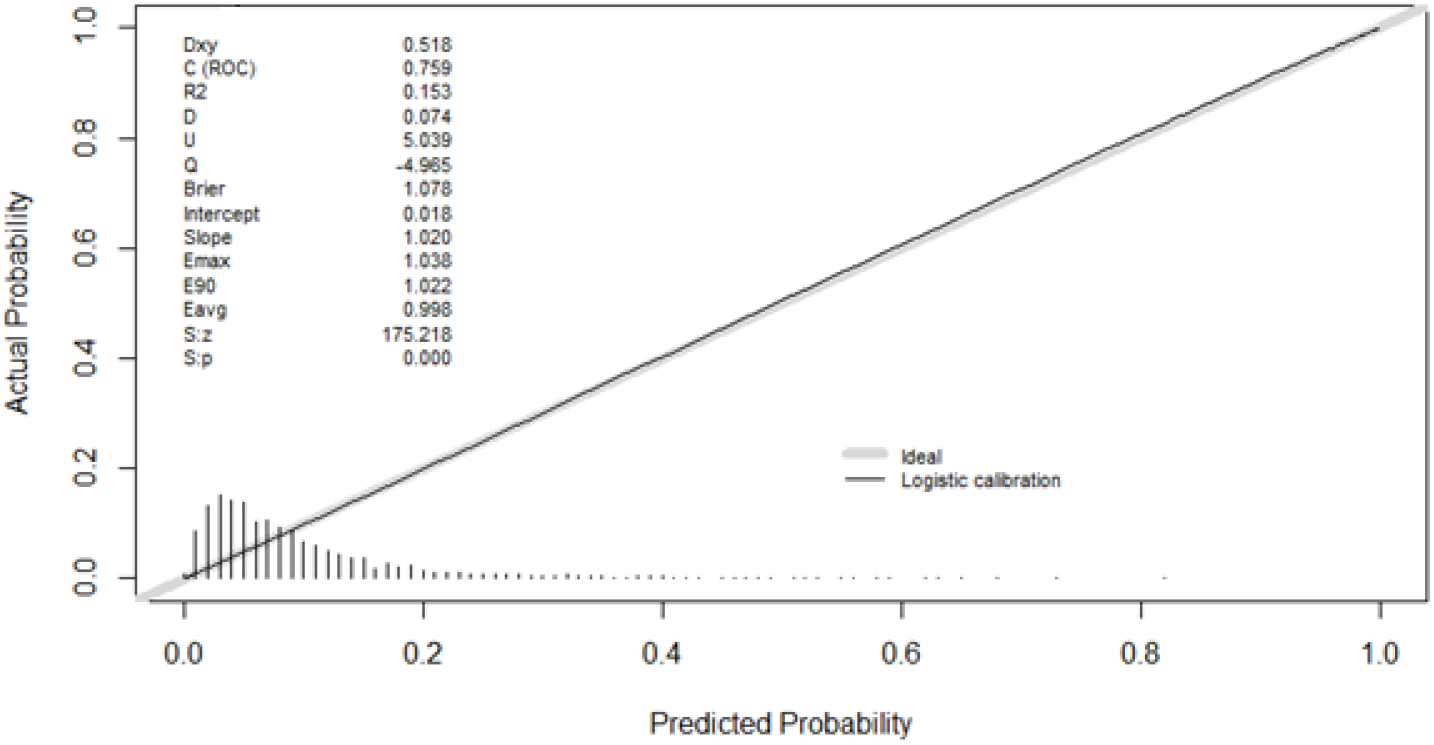
Calibration of HDP combined model tested in BiB.

Figure 5 shows the calibration slope for the combined model for LGA.

**Figure 5.**
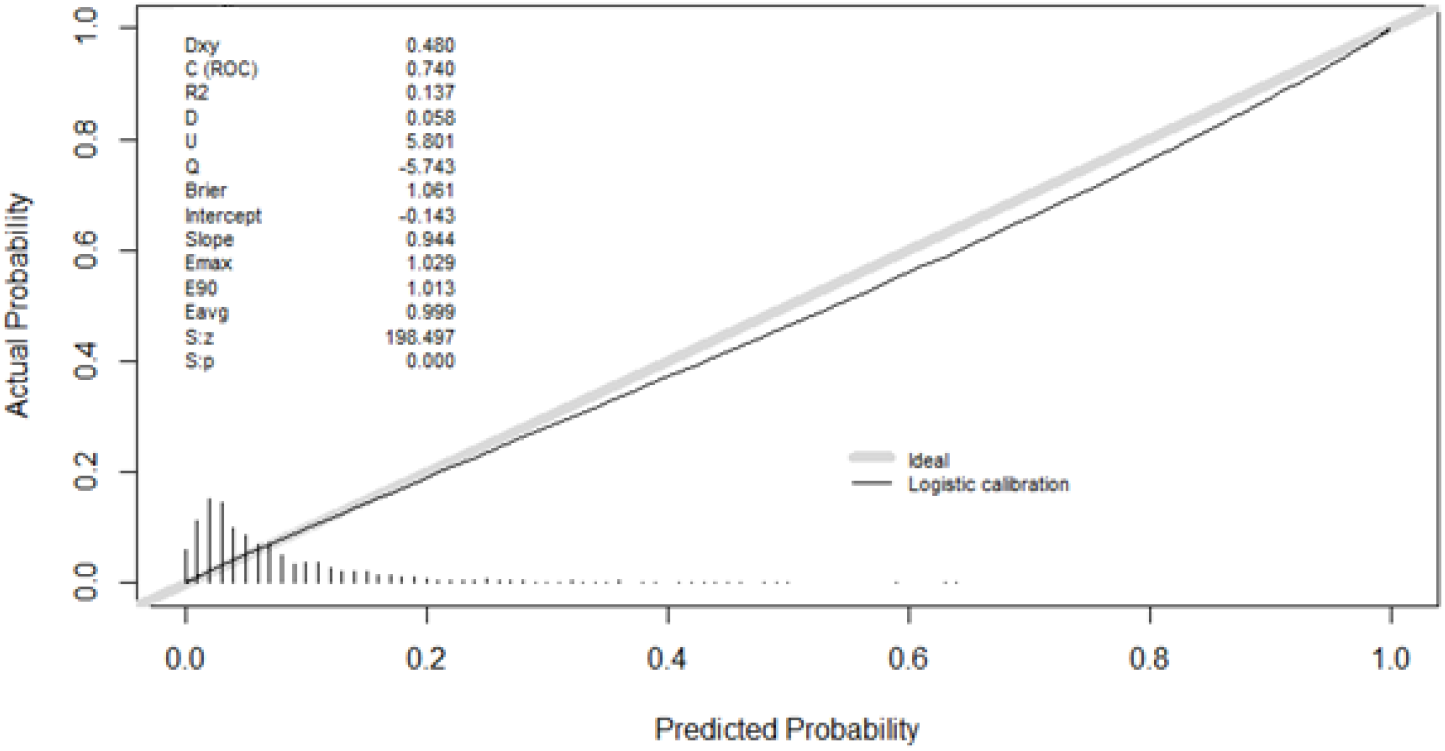
Calibration of LGA combined model tested in BiB

### External validation

External validation in UPBEAT revealed similar patterns of results to those in BiB (Figure 2). AUC was higher for the GDM and HDP combined models when compared to the risk factor models. However, across all models, we saw lower discrimination (AUC lower by ∼1). For example, the combined model AUC (95% CI) for GDM was 0.78 (0.74,0.81) in BiB and 0.62 (0.56,0.69) in UPBEAT. Equivalent results for HDP were AUC (95% CI) 0.76 (0.73,0.79) in BiB and 0.62 (0.55,0.69) in UPBEAT.

### Sensitivity analysis

We did not find that criteria used to diagnose GDM had much influence upon the results. The combined risk factor and metabolite model for the UPBEAT GD models using the IADSPG criteria was AUC (95% CI) 0.64 (0.60,0.68). Using the WHO criteria, as in BiB, the combined model discrimination was AUC (95% CI) 0.65 (0.58,0.71) (Table S6).

The strength and direction of association between BMI and each outcome was similar in the whole BiB cohort and the BiB cohort including only obese women; associations in UPBEAT were weaker than either BiB dataset (Table S7).

To assess the possibility that one predictive model could predict more than one outcome - we evaluated the discrimination of models developed for outcomes for which they were not trained. None of the models performed as well when applied to different outcomes to those (Table S8).

Performances of models in UPBEAT were similar when applied to NMR metabolites obtained from ∼15-week samples (Table S9). The combined model AUC was the same for HDP (AUC 0.62) at both timepoints. The combined model AUC was similar for GDM (AUC (95% CI) 0.62 (0.57,0.66) and 0.65 (0.60,0.69)) at 15 and 27 weeks, respectively), LGA (AUC (95% CI) 0.52 (0.45,0.59) and 0.57 (0.51,0.63), SGA (AUC (95% CI) 0.51(0.43,0.59) and 0.55 (0.47,0.62)) and PTB (AUC (95% CI) 0.52 (0.42,0.62) and 0.54 (0.44,0.64). There was good correlation between the measures at the two timepoints (mean correlation 0.68) (Table S10).

When we trained and tested models for spontaneous PTB in BiB, we obtained a combined model for spontaneous PTB that had better discrimination than any (iatrogenic or spontaneous) PTB. The combined model AUC (95% CI) for spontaneous PTB was 0.58 (0.51,0.65) compared to AUC (95% CI) 0.53 (0.48,0.59) for any PTB. However, the risk factor only model had the highest AUC (95% CI) at 0.65 (0.57,0.72), with the metabolite only model performing poorly (AUC (95% CI) 0.48 (0.42, 0.56)) (Table S6).

## Discussion

Using data from a large multi-ethnic cohort we have shown good discrimination and calibration for GDM, HDP, and LGA can be obtained from a combination of established risk factors and metabolites. The overall pattern of discrimination results was validated in a smaller independent cohort of obese pregnant women, though the AUC’s were weaker. These findings show promise for the use of NMR-derived metabolites to improve prediction of common pregnancy complications, though we acknowledge the need to undertake further validation in a large independent sample of unselected women. To date we have not been able to find such a study.

The proportion of GDM was more than three times greater in UPBEAT compared with BiB when you used the IADSPG criteria. The proportion was more similar, but still higher (10.3% in UPBEAT compared to 8.1% in BiB) when using the WHO criteria. The lower proportion of those who are SGA, and higher proportion who are LGA in UPBEAT is also likely to reflect the fact that UPBEAT includes only obese women. The prevalence of HDP and PTB was similar between the two cohorts.

We found little overlap in the risk factor and metabolite predictors retained in models for each outcome. Risk factors were retained in the combined models for all pregnancy-related disorders. A small number of the metabolites were retained in the prediction models for more than one outcome. Specifically, apolipoproteins, monounsaturated fatty acids and triglycerides were retained in the prediction models for GDM, LGA and SGA.

The overall best discrimination was seen for the combined (established risk factors and metabolite) models for predicting GDM, HDP and LGA. Discrimination for GDM with the combined model (AUC 0.78) was similar to that previously reported for GDM prediction based on clinical information, such as previous history of GDM or LGA, and sociodemographic characteristics (AUC ∼0.78) ^43^. It performs better than a previous reported model of risk factor variables (age, previous GDM, family history of type 2 diabetes, systolic blood pressure, skinfold thicknesses, and waist to height/neck to thigh ratios (AUC 0.71). This risk factor model improved when it included biomarkers such as glucose, adiponectin, sex hormone binding globulin and triglycerides (AUC 0.77), but not with the addition of NMR metabolites (AUC 0.77). ^29^ However, our combined model has the advantage in that it can be applied to nulliparous women and does not rely on personal and family medical history. The combined models for GDM, HDP and LGA in our study had good discrimination and calibration. One aim of this study was to explore the extent to which a group of potential predictors (metabolites or established risk factors) might predict several pregnancy outcomes. However, the best performing models (combined models for GDM, LGA and HDP) showed only modest discrimination for other outcomes (AUC ranging from 0.60 – 0.68), with the strongest being for the prediction of LGA using the GD combined model (Table S8). Overall, these findings for the NMR metabolite platform suggest that it may not be possible to develop a single prediction model that is accurate for several adverse pregnancy outcomes.

For HDP and SGA, whilst the combined models had good discrimination, the metabolites did not substantially improve the discrimination or calibration when compared to the established risk factors. In the interests of maximising the sample, our HDP variable included both gestational hypertension and PE, and our model discrimination for HDP was weaker than that seen for the sFlt-1/PlGF ratio for PE alone ^24^ and that seen for a model including first antenatal clinical characteristics and repeat antenatal blood pressure measurements for PE or gestational hypertension alone (AUC 0.77 - 0.88) ^6^. It would be useful to repeat our analyses in a larger study that had sufficient power to explore the prediction accuracy of metabolites for PE and gestational hypertension separately.

Previous studies have reported better discrimination for SGA using metabolite models than reported in this study. However in those studies, sample sizes were small and they did not attempt external validation or assessment of calibration ^25 26^. We used a <10% cut off for SGA, as recommended by the WHO. Some recommendations advise using a more conservative <3% cut off ^28^, whilst there is also evidence that a threshold of 25% better predicts stillbirth and neonatal mortality^44^. We lacked power in this study to explore a range of different thresholds for SGA and LGA and be able to precisely detect differences between them.

For any PTB (iatrogenic or spontaneous), discrimination was very poor across all models. When we ran the analyses limited to spontaneous PTB, we found the discrimination for all models was higher than that seen for the models with any PTB. However, the AUC remained poor for the combined (AUC (95% CI) 0.58 (0.51,065)) and metabolites alone model (AUC (95% CI) 0.48 (0.41,0.56)), with modest discrimination for the risk factor model (AUC (95% CI) 0.65 (0.57,0.72). We acknowledge that by its very nature, spontaneous PTB is difficult to predict. Our results demonstrate the need for better models to predict PTB, aside from a previous history of PTB. Our results also suggest that metabolomics quantified using the NMR platform are not useful for predicting iatrogenic or spontaneous PTB.

We were unable to identify a general population of pregnant women with relevant data for validation, so we performed validation in obese pregnant women (UPBEAT). In this sample, models demonstrated poorer discrimination. It is expected that prediction is poorer in external validation samples ^45^, but it is also likely that this has also been influenced by the different incidences of some outcomes between the two cohorts and the distinct metabolic perturbations experienced by obese women during pregnancy ^23^.

## Strengths and limitations

Previous studies aiming to improve prediction of pregnancy-related disorders often do not compare performance to established risk factors, assess calibration or undertake external validation as we have done here ^46 47 47 48 49^. BiB has considerably larger numbers of women with NMR data than previous studies of metabolite prediction. The NMR platform has several strengths in relation to its use for prediction; measurements are reliable with little variation between batches, the volume of plasma or serum required for analyses is small (100-300 microlitres) and to obtain all measures is not expensive (∼£20) ^50^. NMR provides absolute quantification, which can represent clinically useful units. However, the platform quantifies only a small proportion of the metabolome. Other platforms, such as Metabolon mass spectrometry, are able to quantify over 1000 metabolites ^51^. With greater coverage of the metabolome it is possible that we would have improved prediction for the pregnancy outcomes explored here. We were limited in this study by the BiB NMR samples being taken in the second trimester. However, when we performed the validation using the 15-week gestation data from UPBEAT, the results were comparable to second trimester results in UPBEAT (Table S9) and metabolites at 26 weeks correlate with those at 15-weeks (Table S10). Taken together, these suggest that the metabolites measured in the second trimester are good proxies for earlier antenatal measures of the same metabolites. However, this needs to be directly tested. Ideally, we would have a prediction tool that could be used as early as possible in pregnancy. It would be able to be repeated so that women’s antenatal care could be tailored to their risk from early pregnancy and updated with repeat assessment if risk changed.

## Concluding remarks

To conclude, our results suggest metabolomics combined with established risk factors improve prediction of GD, HDP and LGA, compared to established risk factors alone. As we were only able to explore validation in a select cohort of obese women, we need to validate these findings in large, general cohorts of pregnant women. A predictive test for all or several of these outcomes would have significant clinical importance and allow us to identify mothers in need of further resources and antenatal monitoring. However, we found relatively little overlap in the models for different outcomes and poor discrimination for other outcomes for any combined model than the outcome it had been developed for. By improving the allocation of resources and stratifying antenatal care from early pregnancy until delivery, we can reduce the burden on the healthcare providers and the morbidity and mortality of mothers and offspring.

## Data Availability

Data are available from the studies used in this paper upon request: Born in Bradford (BiB) and the UK Pregnancies Better Eating Activity Trial (UPBEAT).

https://borninbradford.nhs.uk/

https://www.ucl.ac.uk/epidemiology-health-care/research/bsh/research/obesity/uk-pregnancies-better-eating-and-activity-trial-upbeat

## Acknowledgments

The authors are extremely grateful to all the families who took part in this study, and the teams that make up BiB and UPBEAT, which includes midwives, interviewers, computer and laboratory technicians, clerical workers, research scientists, volunteers, managers, receptionists and nurses.

## Abbreviations

GDM: gestational diabetes mellitus
HDP: hypertensive disorders of pregnancy
SGA: small for gestational age
LGA: large for gestational age
PTB: preterm birth
BMI: body mass index
BiB: Born in Bradford
UPBEAT: UK Pregnancies and Better Eating Activity Trial
AUC: Area under the curve
NMR: nuclear magnetic resonance

